# Tixagevimab-cilgavimab as an early treatment for COVID-19 in kidney transplant recipients

**DOI:** 10.1101/2022.09.30.22280568

**Authors:** Ilies Benotmane, Jérôme Olagne, Gabriela Gautier Vargas, Noëlle Cognard, Francoise Heibel, Laura Braun-Parvez, Nicolas Keller, Jonas Martzloff, Peggy Perrin, Romain Pszczolinski, Bruno Moulin, Samira Fafi-Kremer, Sophie Caillard

## Abstract

**Objective:** This single-center retrospective study evaluated the use of tixagevimab-cilgavimab as an early treatment for COVID-19 in kidney transplant recipients (KTRs) during the omicron wave.

**Methods:** KTRs were deemed at high risk for moderate-to-severe COVID-19 in presence of at least one comorbidity (age >60 years, diabetes, obesity, or cardiovascular disease) associated with a weak humoral response (<264 BAU/mL). All other KTRs were considered at low risk. The two groups were stratified according to the administration of tixagevimab-cilgavimab and compared in terms of COVID-19-related hospitalization, oxygen need, ICU admission, and mortality.

**Results:** Of the 61 KTRs at high risk, 26 received tixagevimab-cilgavimab. COVID-19-related hospitalizations (3.8% *versus* 34%, p=0.006) and oxygen need (3.8% *versus* 23%, p=0.04) were significantly less frequent in patients who received tixagevimab-cilgavimab. In addition, non-significant trends towards a lower number of ICU admissions (3.8% *versus* 14.3% p=0.17) and deaths (0 *versus* 3, p=0.13) were observed after administration of tixagevimab-cilgavimab. Ten of the 73 low-risk KTRs received tixagevimab-cilgavimab, and no significant clinical benefit was observed in this subgroup.

**Conclusion:** Early administration of tixagevimab-cilgavimab may be clinically useful in high-risk KTRs with COVID-19; however, no major benefit was observed for low-risk patients.

---

The transmissibility of Omicron is markedly higher than that of prior SARS-CoV-2 variants – possibly because of greater immune escape after vaccination and/or natural infection. However, this is counterbalanced by less severe clinical manifestations in the general population.^1^ Unfortunately, immunocompromised kidney transplant recipients (KTRs) infected by Omicron are still at risk of developing severe COVID-19 – possibly because of their weak vaccine response.^2,3^ Notably, Omicron has a higher ability than other variants to escape vaccine-induced humoral immune responses.^4^ In addition, there have been reports of a greater mortality in transplanted patients than in general population during the Omicron wave.^5–7^

Anti-SARS-CoV-2 monoclonal antibodies (mAbs) have been successfully used for pre- and post-exposure prophylaxis and as an early treatment for patients at high risk of developing severe COVID-19 – including KTRs.^8–12^ Currently, their administration is the mainstay of COVID-19 management in solid organ transplant recipients who did not develop a sufficient humoral immune response following vaccination. Accordingly, remdesivir has shown limited clinical benefits in this population^13^ and Paxlovid^®^ administration may be problematic because of its interactions with mTOR and/or calcineurin inhibitors.^14^ Unfortunately, the Omicron variant and its sublineages are capable of escaping the vast majority of currently available mAbs – and especially the casirivimab-imdevimab combination.^15,16^ As for other options, sotrovimab is effective against BA.1 sublineages but it has been rapidly abandoned since the emergence of the BA.2, BA.4, and BA.5 breakthroughs.^17^ Bebtelovimab is clinically effective^18^ but its availability is currently limited to the United States.^19^ The tixagevimab-cilgavimab combination retains a neutralizing activity against all of the Omicron sublineages – albeit at a lesser extent than that observed against prior variants.^17,20–22^ While tixagevimab-cilgavimab has been originally developed for pre-exposure prophylaxis,^8^ it represented the only efficient mAbs combination available in France during the Omicron wave. In this scenario, French health authorities have granted authorization for its use for both post-exposure prophylaxis and as an early treatment in patients deemed at high risk of developing moderate-to-severe COVID-19.^23^ This single-center retrospective study describes the use of tixagevimab-cilgavimab as an early treatment for COVID-19 in KTRs during the Omicron wave.

## Patients and Methods

The study protocol was approved by the local Ethics Committee (comité de protection des personnes de Strasbourg, France, identifier: DC-2013–1990) and written informed consent was obtained from all participants. As of December 2021, Omicron has been the predominant SARS-CoV-2 variant in France. The study sample consisted of all adult KTRs with COVID-19 who were followed in the outpatient clinic of the Department of Nephrology, Dialysis, and Transplantation, Strasbourg Hospital University (Strasbourg, France), between December 22, 2021 and April 27, 2022. All KTRs were instructed to contact the Department in the event of COVID-19 infection – which was diagnosed in presence of positive reverse transcriptase-polymerase chain reaction (RT-PCR) results on nasopharyngeal swab specimens. Tixagevimab-cilgavimab was used in our center as of the emergence of BA.1 and BA.2 breakthroughs. Adult patients received a single dose of 600 mg (300 mg of each antibody) given intravenously.

According to the recommendations set forth by French health authorities, KTRs with confirmed or suspected infection caused by the Omicron variant were eligible to receive tixagevimab-cilgavimab with curative intent. Based on the presence of comorbidities and the extent of serological protection,^24^ the study participants were retrospectively divided into two risk groups (high *versus* low risk) for developing moderate-to-severe COVID-19. Patients with at least one comorbid condition (i.e., age > 60 years, diabetes, obesity, or a history of cardiovascular disease) who were unvaccinated or showed a weak vaccine-induced humoral immune response (< 264 binding antibody units [BAU]/mL) were considered at high risk. All other patients (i.e., those without comorbidities or those who showed a satisfactory humoral immune response defined by an antibody titer ≥264 BAU/mL) were deemed at low risk. KTRs who received tixagevimab-cilgavimab as an early treatment were compared with those who did not in terms of COVID-19-related hospitalization, oxygen need, intensive care unit (ICU) admission, and mortality. Patients who were treated after more than 8 days from the appearance of symptoms and those with rapid onset of severe symptoms requiring immediate hospitalization were excluded.

Continuous data are given as means and standard deviations and analyzed using the Welch test or the Mann-Whitney *U* test, as appropriate. Categorical variables are expressed as counts and percentages and compared using the chi-square test or the Fisher’s exact test, as appropriate. The Kaplan-Meier method (log-rank test) was used for intergroup comparisons of COVID-19-related hospitalizations, oxygen need, ICU admissions, and mortality. All calculations were undertaken in GraphPad Prism, version 8.0 (GraphPad Inc., San Diego, CA, USA) and pvalue.io. Two-tailed p values < .05 were considered statistically significant.

## Results

A total of 197 KTRs with COVID-19 were identified throughout the study period. Of them, 63 were excluded (Figure 1) for the following reasons: treatment with sotrovimab at the beginning of the Omicron wave, n = 31; treatment with mAbs as a late post-hospitalization rescue therapy, n = 17; and lack of serology data to perform risk stratification, n = 15. Of the remaining 134 KTRs, 61 were considered at high risk and 73 at low risk for moderate-to-severe COVID-19, respectively. Finally, 36 KTRs received tixagevimab-cilgavimab within an early curative scheme. No adverse effect was observed in any patient.

**Figure 1.**
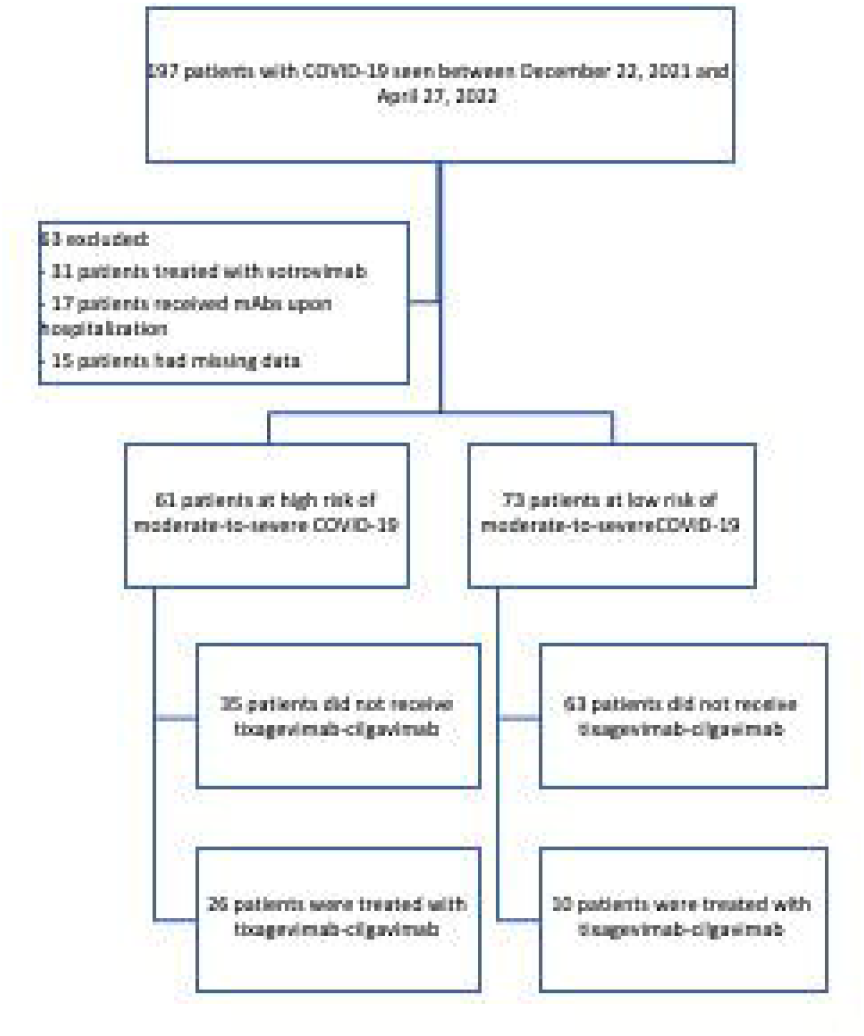
Flow of patients through the study. * Patients who harbored at least one comorbid condition (i.e., age > 60 years, diabetes, obesity, or a history of cardiovascular disease) and those unvaccinated or with a weak vaccine-induced humoral immune response (< 264 binding antibody units [BAU]/mL) were considered at high risk. **Patients without comorbidities who showed a satisfactory humoral immune response against SARS-CoV-2 were considered at low risk.

### Tixagevimab-cilgavimab as an early treatment in high-risk kidney transplant recipients

Of the 61 KTRs at high risk, 57 (93%) have been previously vaccinated against SARS-CoV-2 and 46% had already received tixagevimab-cilgavimab (300 mg) for pre-exposure prophylaxis. The vast majority of KTRs included in this group (n = 55, 92%) developed symptomatic COVID-19. A total of 26 high-risk KTRs received tixagevimab-cilgavimab as an early treatment within a median of 3 days (range: 2−4.75) from the onset of symptoms. The remaining 35 patients were not treated with tixagevimab-cilgavimab for the following reasons: late notification of diagnosis (n = 19), clinical management in a different hospital (n = 4), COVID-19 infection prior to tixagevimab-cilgavimab availability (n = 7), patient refusal (n = 2), and unknown reasons (n = 3). The general characteristics of high-risk KTRs are summarized in Table 1. Compared with untreated patients, those who received tixagevimab-cilgavimab were more likely to have a positive history of cardiovascular disease, had a longer interval from transplantation, and more frequently benefitted from early reduction of immunosuppressive therapy. On analyzing clinical outcomes, both COVID-19-related hospitalizations (3.8% *versus* 34%, respectively, p = 0.006; Figure 2a) and oxygen need (3.8% *versus* 23%, respectively, p = 0.04; Figure 2b) were significantly less frequent in KTRs who were treated with tixagevimab-cilgavimab. Similar, albeit not significant, trends were observed for ICU admissions (3.8% *versus* 14.3%, respectively, p = 0.17; Figure 2c) and mortality (0 *versus* 3, respectively, p = 0.13; Figure 2d).

**Table 1.**
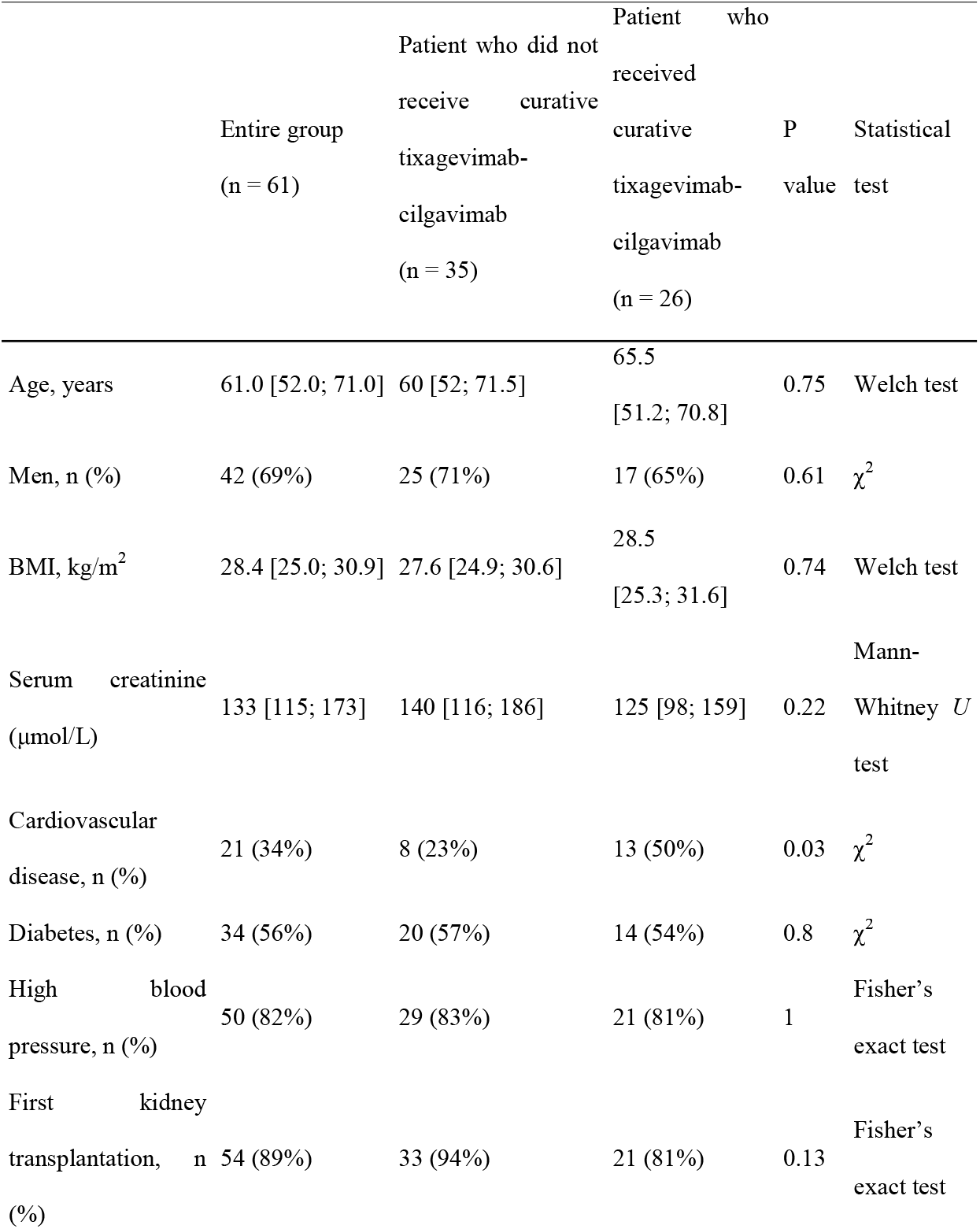
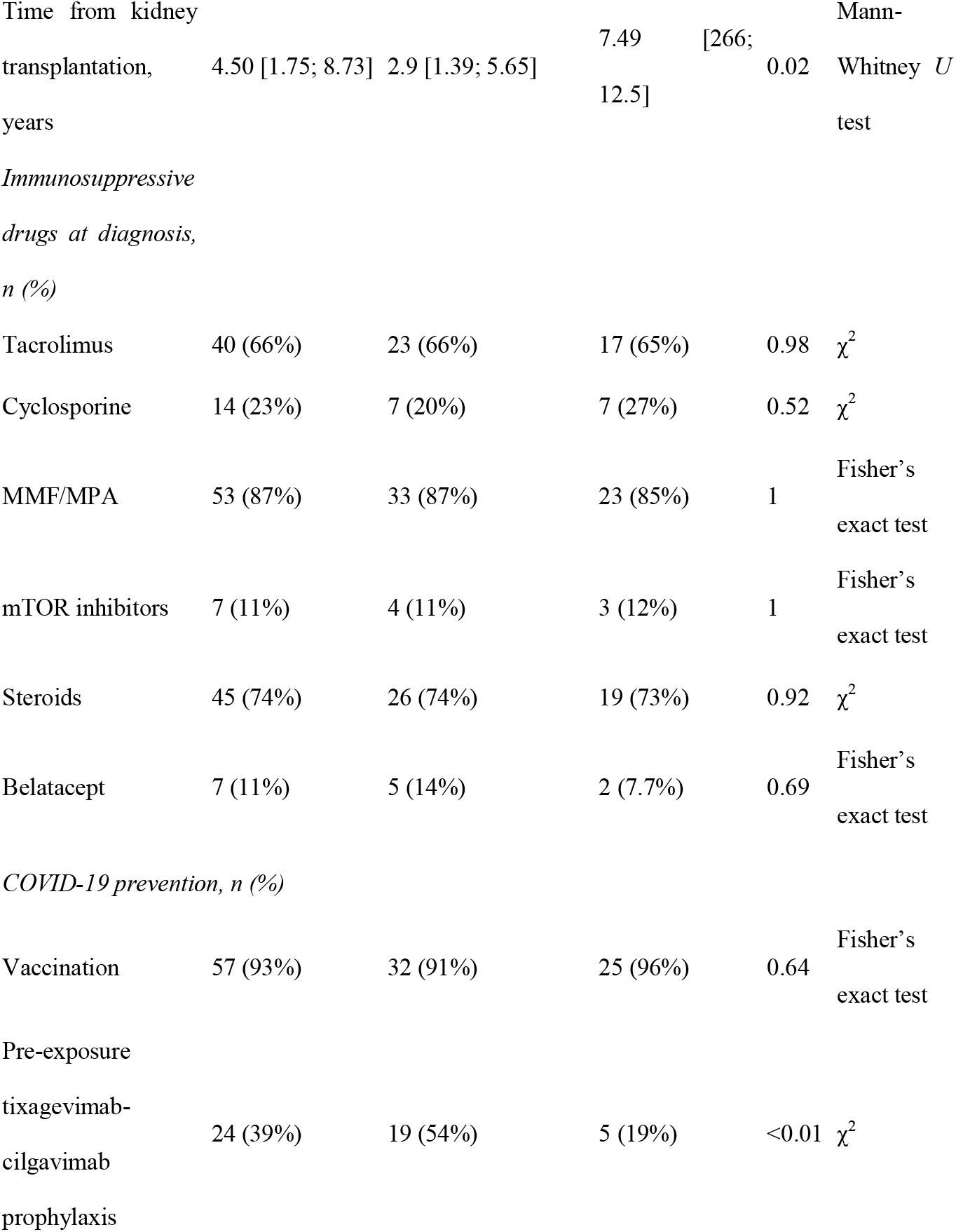
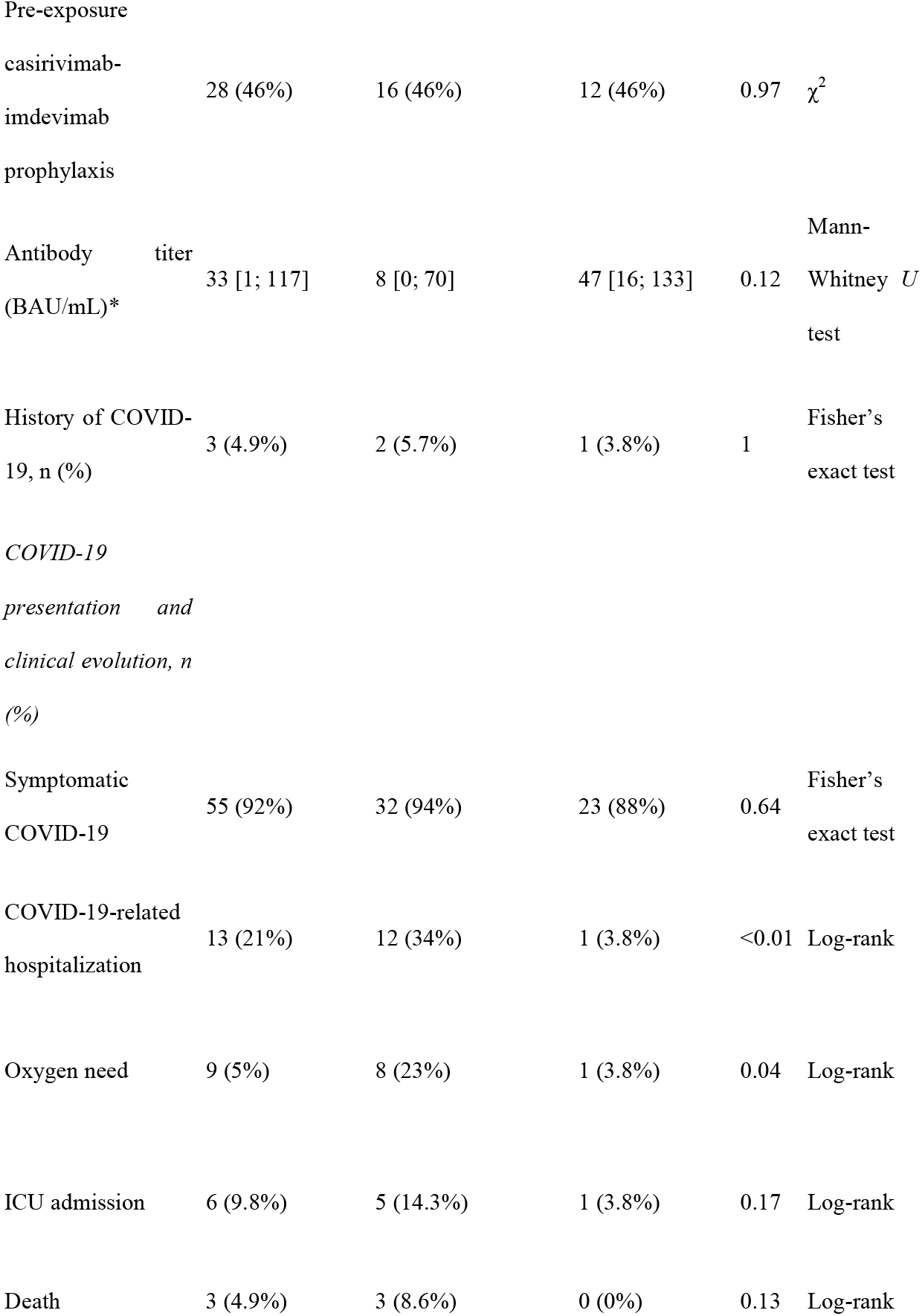
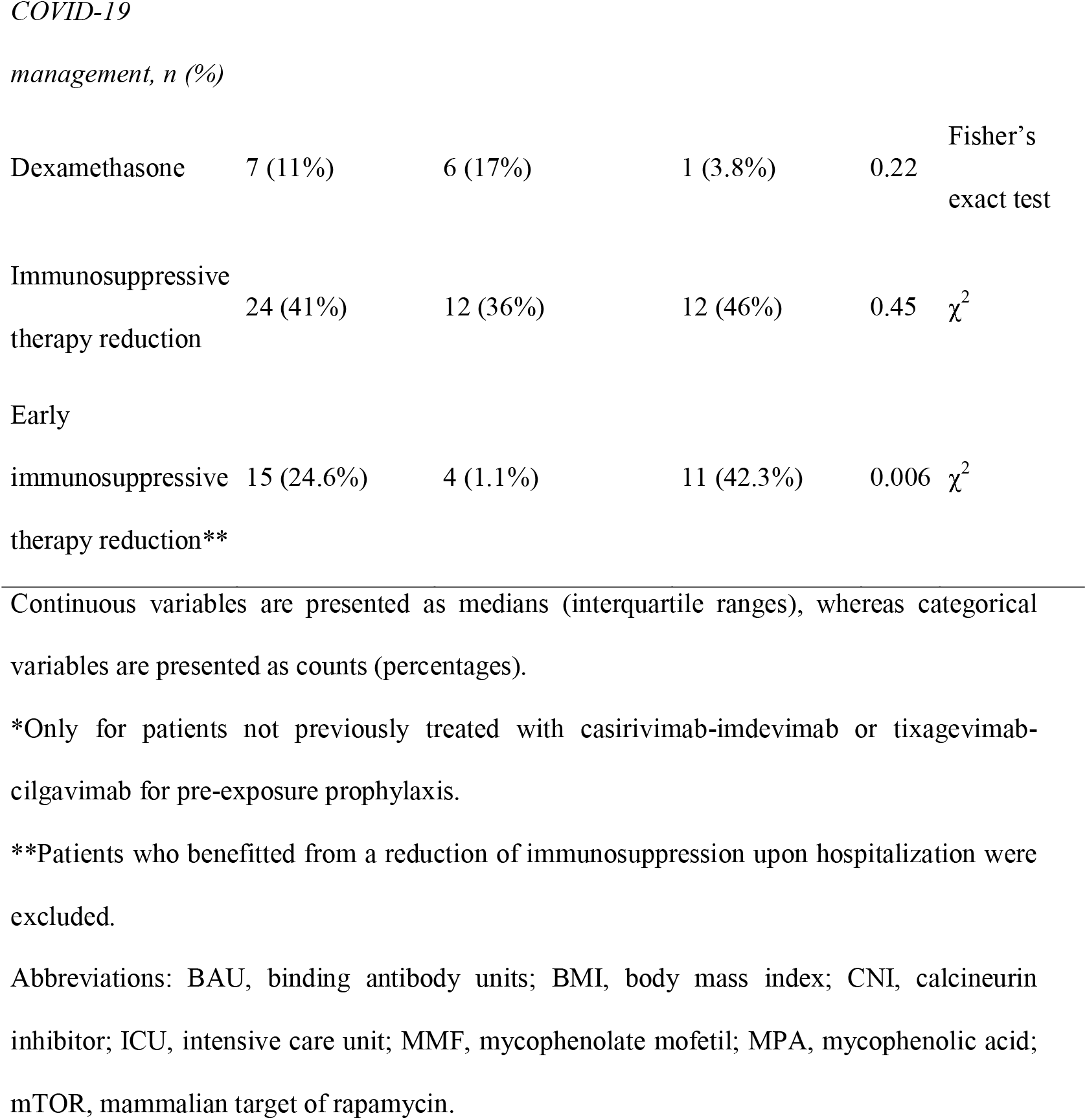
General characteristics of kidney transplant recipients at high risk of moderate-to-severe COVID-19 according to early treatment with tixagevimab-cilgavimab

**Figure 2.**
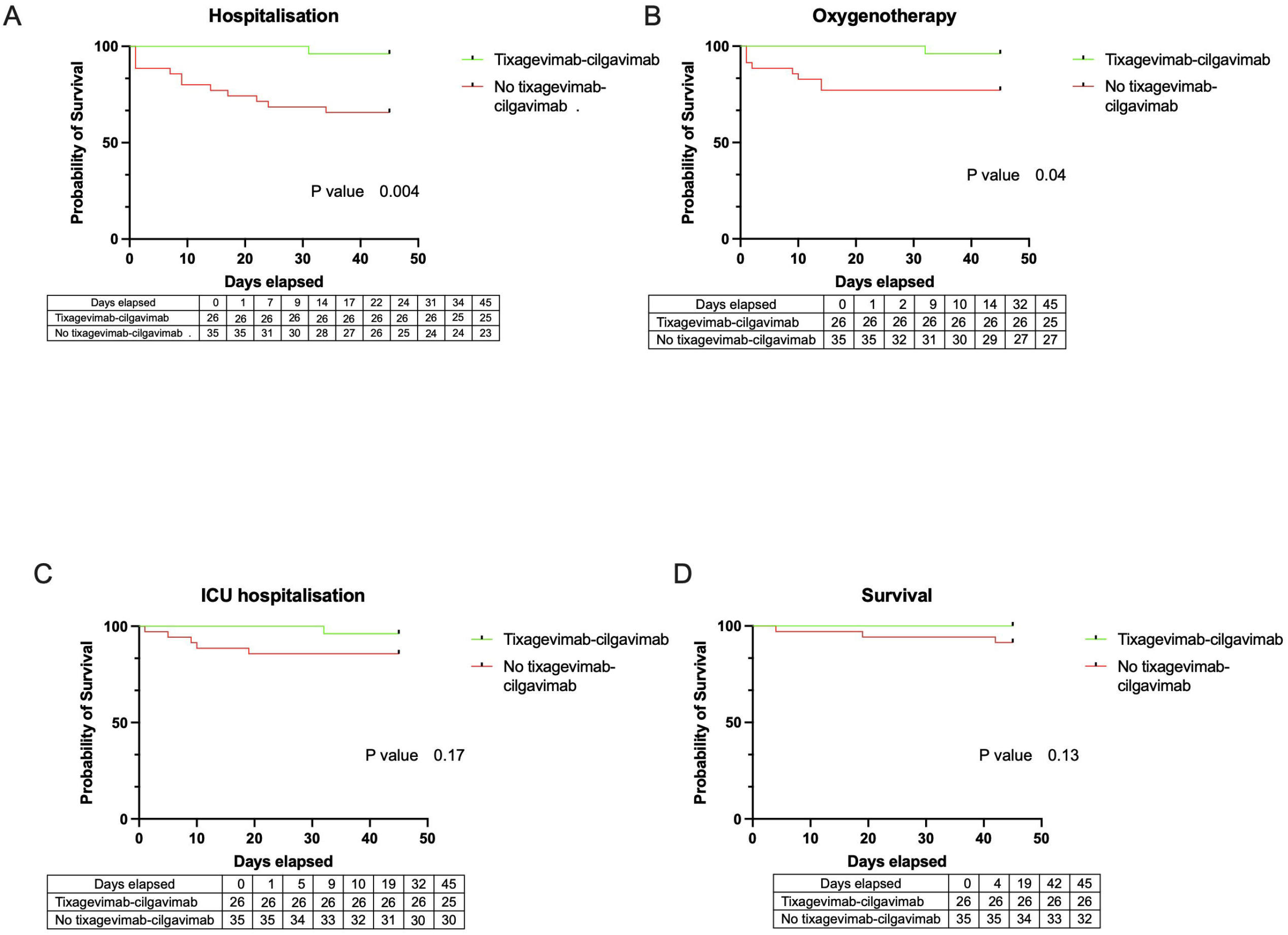
Kaplan-Meier plots of hospitalization-free survival (panel A), oxygen need-free survival (panel B), intensive care unit (ICU) admission-free survival (panel C), and survival (panel D) in high-risk patients stratified according to the use (yes *versus* no) of tixagevimab-cilgavimab as an early treatment for COVID-19.

### Tixagevimab-cilgavimab as an early treatment in low-risk kidney transplant recipients

Of the 73 KTRs at low risk, 69 (95%) have been previously vaccinated against SARS-CoV-2 and 9.6% had already received tixagevimab-cilgavimab (300 mg) for pre-exposure prophylaxis. The majority of KTRs included in this group (n = 58, 88%) developed symptomatic COVID-19. The general characteristics of low-risk KTRs are shown in Table 2. Ten patients received tixagevimab-cilgavimab as an early treatment. Of the 73 KTRs at low risk, only three were hospitalized and one required oxygen treatment. Notably, this KTR received tixagevimab-cilgavimab. No patient required ICU admission or showed COVID-19-related mortality. Low-risk KTRs who received tixagevimab-cilgavimab as an early treatment did not differ significantly from those who did not in terms of COVID-19-related hospitalizations, ICU admissions, and mortality (Table 2 and Figure 3).

**Table 2.**
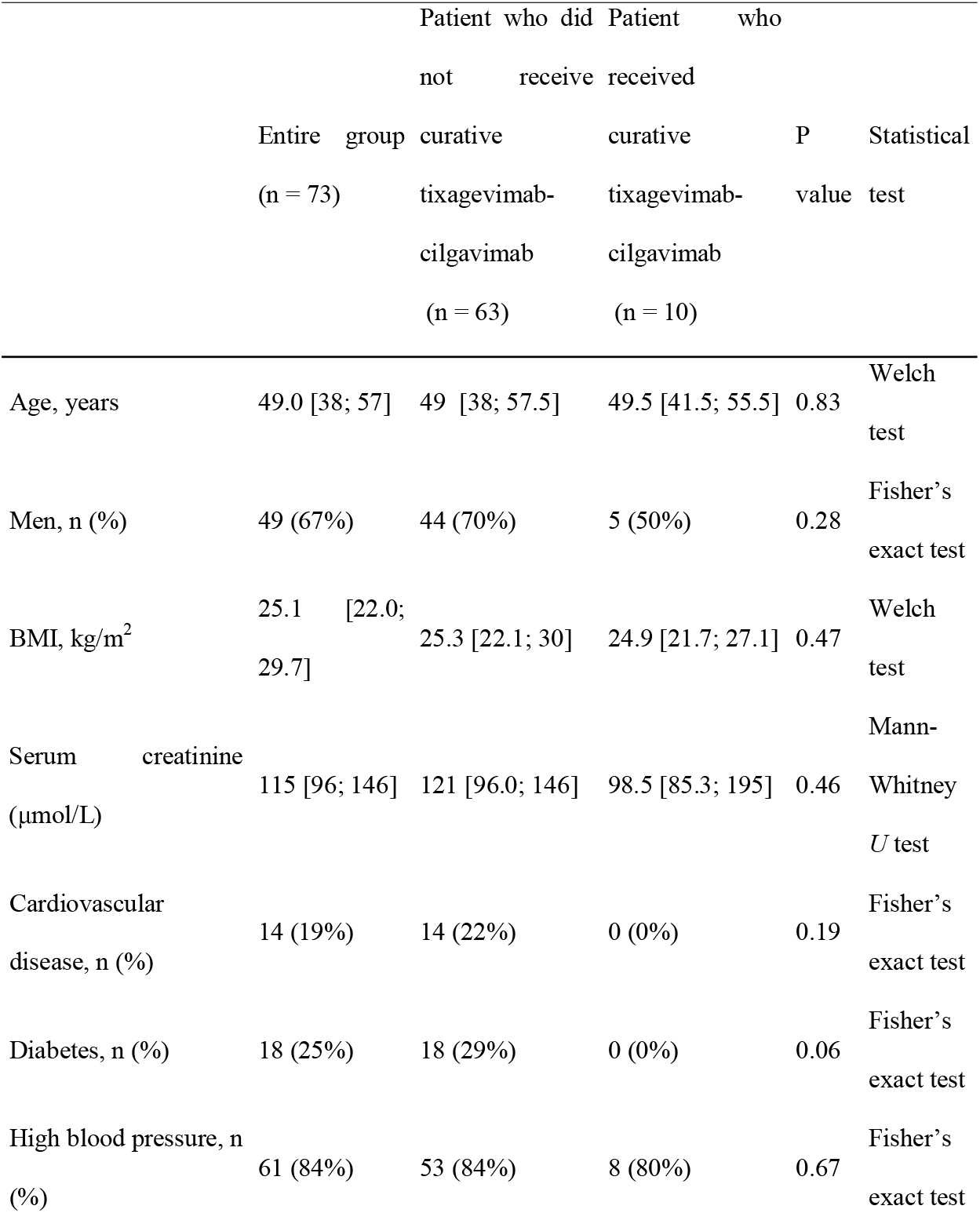
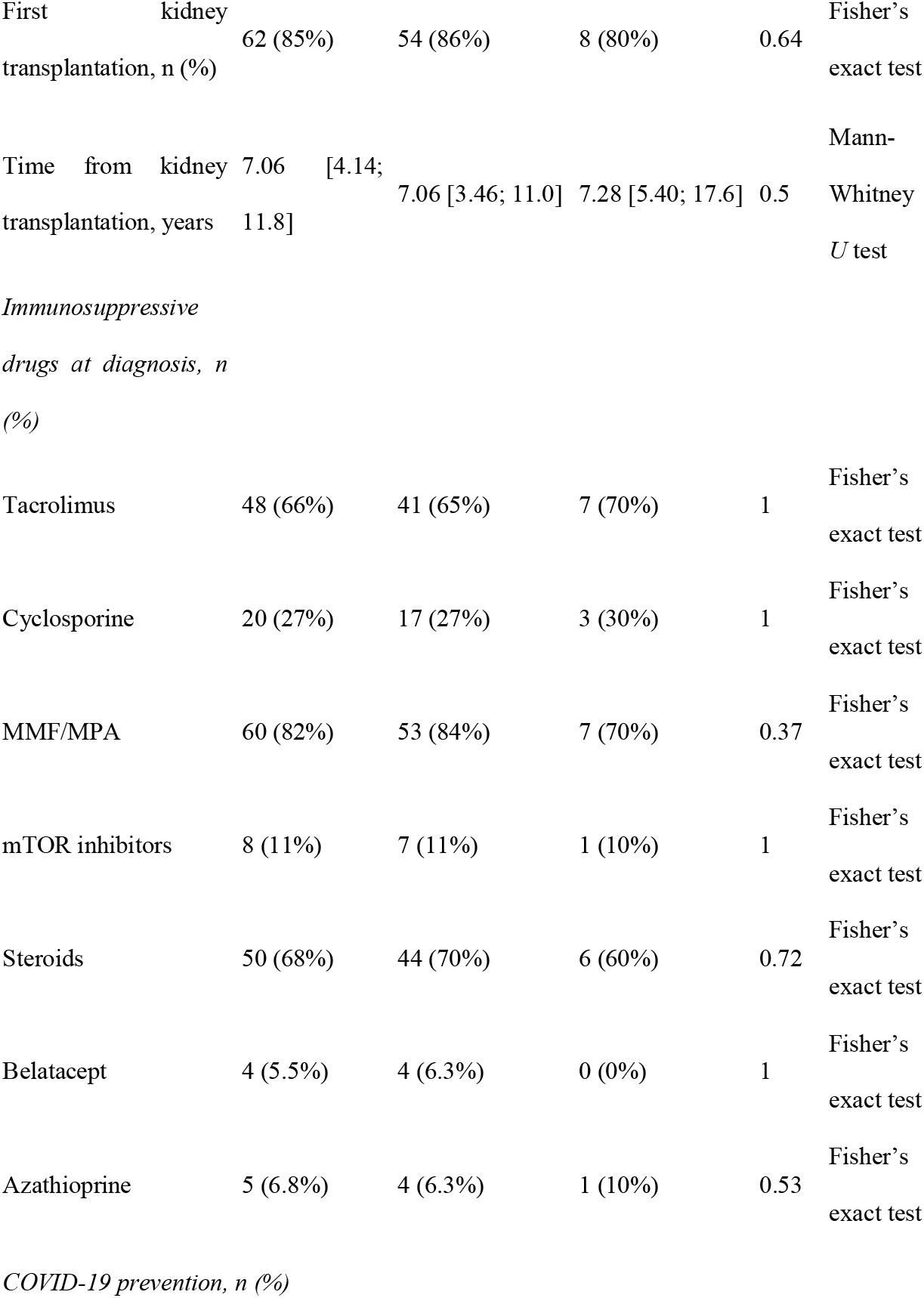
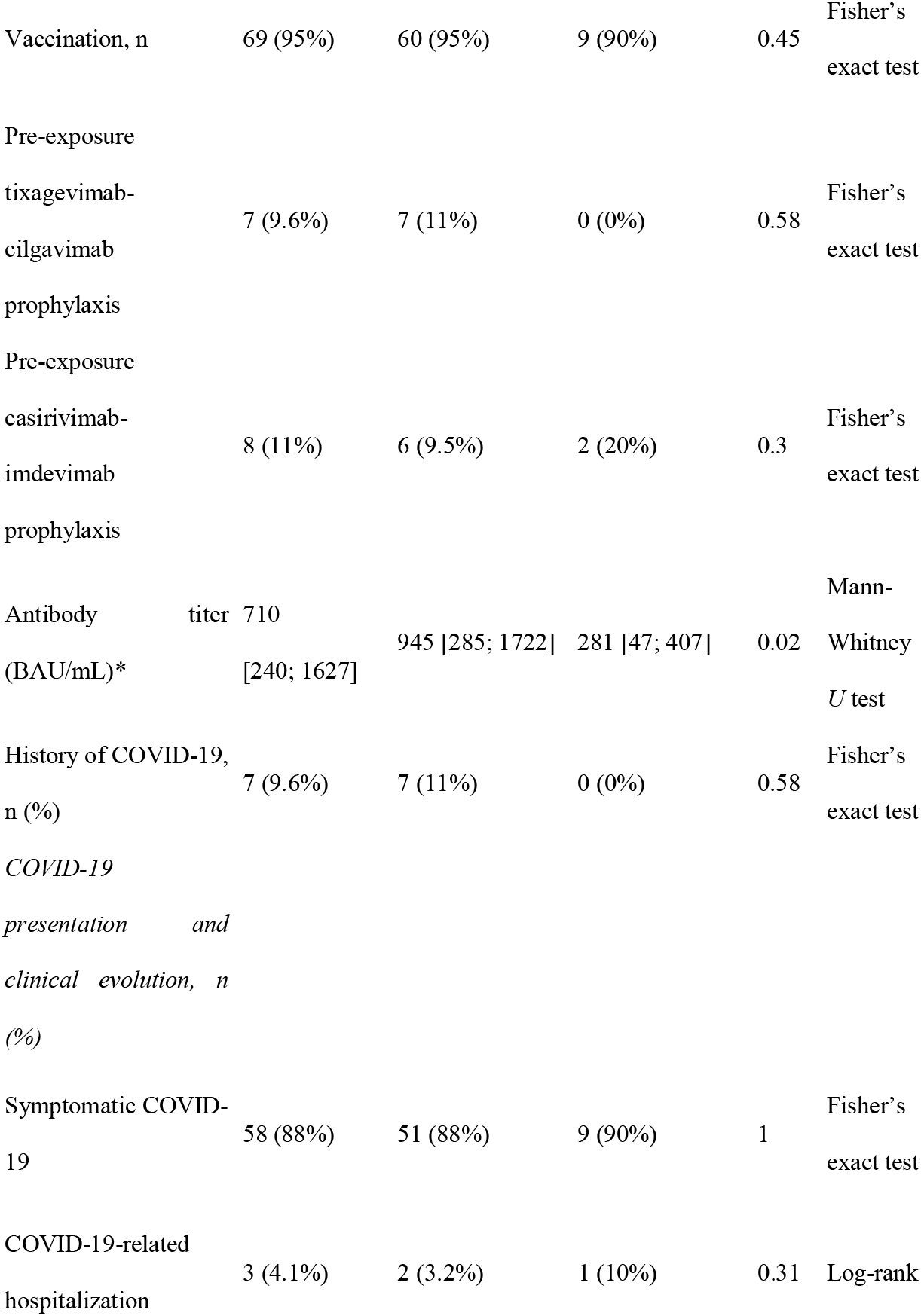
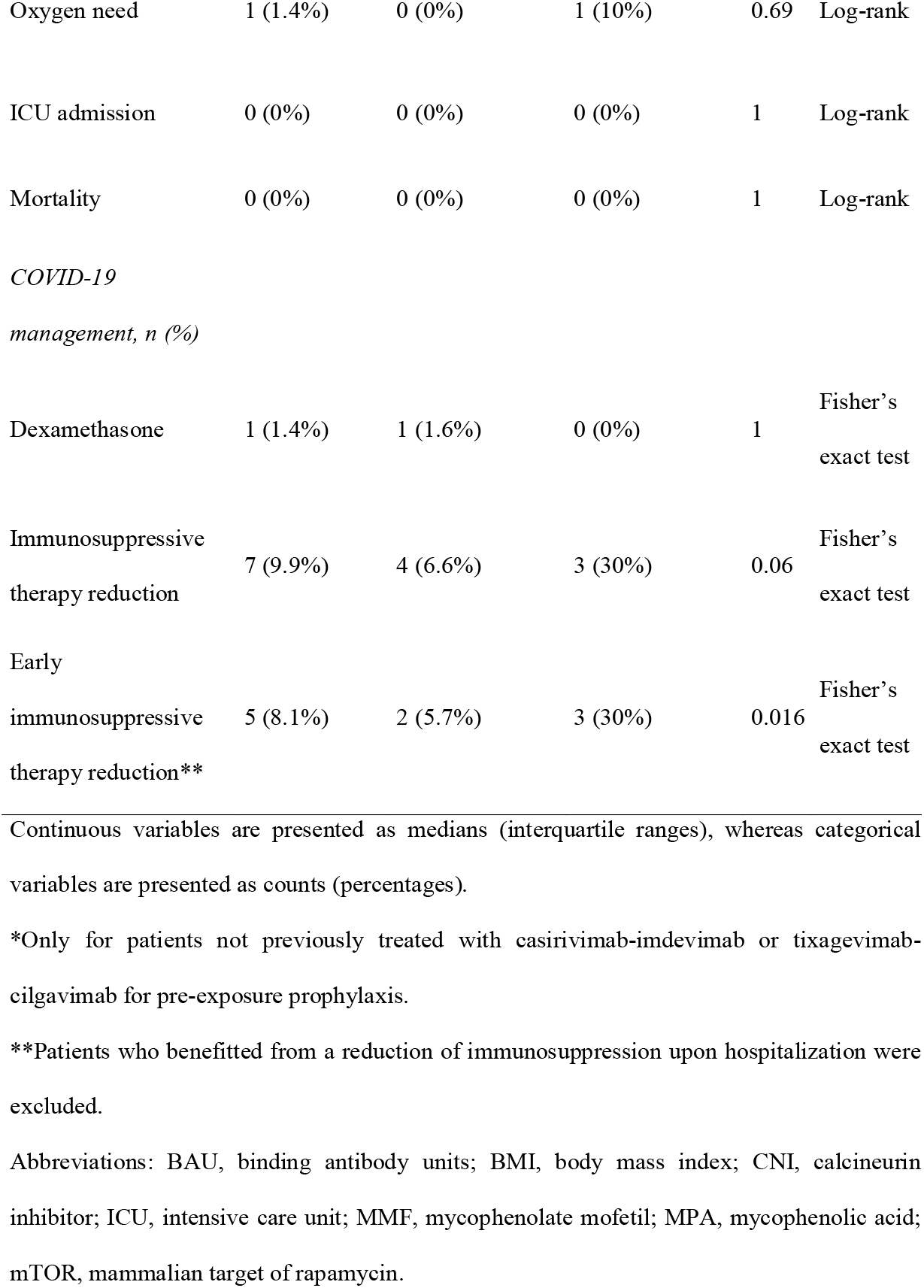
General characteristics of kidney transplant recipients at low risk of moderate-to-severe COVID-19 according to early treatment with tixagevimab-cilgavimab

**Figure 3.**
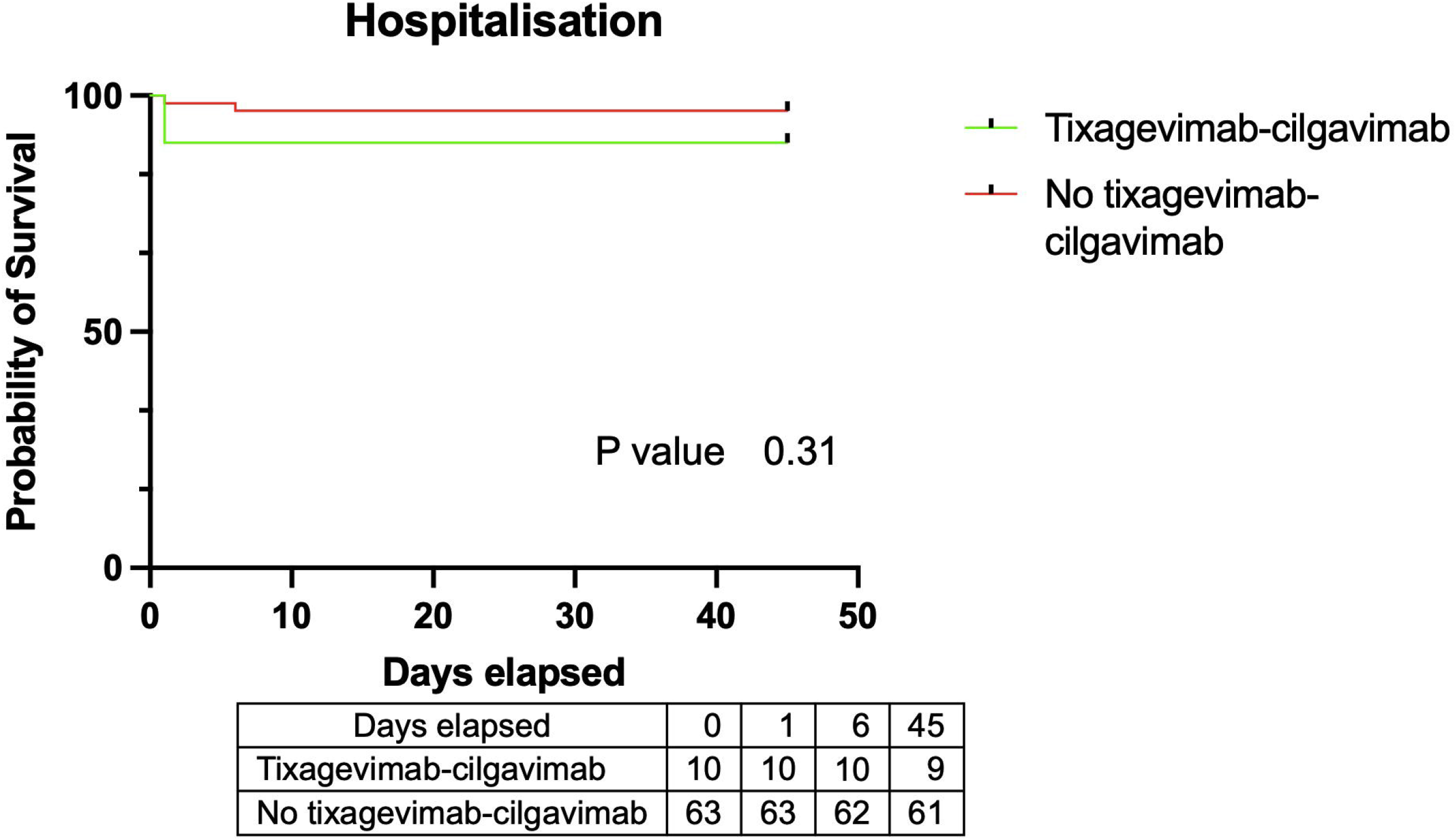
Kaplan-Meier plots of hospitalization-free survival in low-risk patients stratified according to the use (yes *versus* no) of tixagevimab-cilgavimab as an early treatment for COVID-19.

## Discussion

This is, to our knowledge, the first retrospective study that reports the use of tixagevimab-cilgavimab as an early treatment for COVID-19 in an immunocompromised population of KTRs during the Omicron wave. On the one hand, our results indicated that tixagevimab-cilgavimab administration significantly decreased COVID-19-related hospitalizations in KTRs deemed at high-risk for moderate-to-severe COVID-19; notably, this effect was accompanied by non-significant trends towards lower ICU admissions, oxygen need, and mortality. On the other hand, low-risk KTRs did not experience major clinical benefits from tixagevimab-cilgavimab administration as an early curative treatment. In a double-blind, randomized controlled trial, Montgomery et al.^25^ have previously reported a 50% reduction of severe disease and COVID-19-related mortality in patients with documented SARS-CoV-2 infection who received tixagevimab-cilgavimab within 7 days of symptom onset. Unfortunately, this study was conducted in unvaccinated patients and prior to the emergence of Omicron.^25^

Although tixagevimab-cilgavimab was well-tolerated in our cohort, this combination should nonetheless be used cautiously. Vellas et al. have recently shown that tixagevimab-cilgavimab may lead to the selection of cilgavimab-resistant mutants – possibly because of a reduced activity of this combination against the Omicron variant.^26^ Based on our current results, we recommend limiting the administration of tixagevimab-cilgavimab as an early treatment only in KTRs at high risk of developing moderate-to-severe disease. In addition, a thorough virologic monitoring is recommended to identify the emergence of resistant strains.

Our current results should be interpreted in light of some caveats. This is a retrospective, single-center study without a randomized treatment allocation. In addition, the sample size was relatively limited. Despite these limitations, our study shows that, during the Omicron wave, early administration of tixagevimab-cilgavimab associated with reduction immunosuppressive may be clinically useful in high-risk KTRs with COVID-19. However, no major benefit was observed for low-risk patients.

While our results should be confirmed in larger prospective randomized studies, the tixagevimab-cilgavimab combination should be given cautiously due to the potential emergence of mutations conferring drug resistance. The development of mAbs specifically designed against the Omicron variant is eagerly awaited in the next future.

## Data Availability

All data produced in the present study are available upon reasonable request to the authors

## Acknowledgements

The authors wish to thank the clinical research team (Danielle Roy, Annie Menguy, and Fanny Hulstein) and the nursing staff (Sandra Ludwiller, Lucile Steinmetz, Christelle Appenzeller, and Fanny Petitjean) for their contribution to the study.

## Data availability statement

Data supporting the findings from this study are available from the corresponding author upon reasonable request.

